# MIAI: Maturation of Immunity Against Influenza - a German birth cohort study with focus on the development of immunity against respiratory viral infections

**DOI:** 10.1101/2024.06.04.24308438

**Authors:** Carina R. Hartmann, Robin Khan, Jennifer Schöning, Maximilian Richter, Maike Willers, Sabine Pirr, Julia Heckmann, Johannes Dirks, Henner Morbach, Monika Konrad, Elena Fries, Magdalene Winkler, Johanna Büchel, Silvia Seidenspinner, Jonas Fischer, Claudia Vollmuth, Martin Meinhardt, Janina Marissen, Mirco Schmolke, Sibylle Haid, Thomas Pietschmann, Simone Backes, Lars Dölken, Ulrike Löber, Thomas Keil, Peter U. Heuschmann, Achim Wöckel, Sagar, Thomas Ulas, Sofia K. Forslund-Startceva, Christoph Härtel, Dorothee Viemann

## Abstract

Respiratory viral infections (RVIs) are a major global contributor to morbidity and mortality. The susceptibility and outcome of RVIs are strongly age-dependent and show considerable inter-population differences, pointing to genetically and/or environmentally driven developmental variability. The factors determining the age-dependency and shaping the age-related changes of human anti-RVI immunity after birth are still elusive. We are conducting a prospective birth cohort study aiming at identifying endogenous and environmental factors associated with the susceptibility to RVIs and their impact on cellular and humoral immune responses against the influenza A virus (IAV), respiratory syncytial virus (RSV) and severe acute respiratory syndrome coronavirus 2 (SARS-CoV-2). The MIAI birth cohort enrolls healthy, full-term neonates born at the University Hospital Würzburg, Germany, with follow-up at four defined time-points during the first year of life. At each study visit, clinical metadata including diet, lifestyle exposures, sociodemographic information, and physical examinations, are collected along with extensive biomaterial sampling. Biomaterials are used to generate comprehensive, integrated multi-omics datasets including transcriptomic, epigenomic, proteomic, metabolomic and microbiomic methods. The results are expected to capture a holistic picture of the variability of immune trajectories with a focus on cellular and humoral key players involved in the defense of RVIs and the impact of host and environmental factors thereon. Thereby, MIAI aims at providing insights that allow unraveling molecular mechanisms that can be targeted to promote the development of competent anti-RVI immunity in early life and prevent severe RVIs.

**Trial registration number:** DRKS00034278

## INTRODUCTION

Several epidemiological studies demonstrate strong age-dependent differences for the susceptibility and outcome of respiratory viral infections (RVIs) (1). Prime examples are infections with the respiratory syncytial virus (RSV), the influenza A virus (IAV) and the severe acute respiratory syndrome coronavirus 2 (SARS-CoV-2).

RSV is the most common cause of severe respiratory infection in infants, leading to over 3 million hospitalizations and around 66,000 deaths worldwide each year (2–4). Remarkable age-related differences in innate cytokine responses following recognition of RSV have been observed (3, 5), suggesting a critical role of the host response in the disease pathology and clinical outcome. RSV infects virtually all children by the age of three years and then repeatedly infects throughout life. Therefore, host factors of the epithelial barrier and the innate immune system might define the susceptibility to RSV infections, while adaptive immunity, which underlies profound maturational changes during early life, might rather contribute to the severity of the course of an established RSV infections but does not necessarily acquire long-lasting protective memory.

In contrast to the age profile of RSV infections, work from the Global Burden of Disease Study showed that the population attributable fraction of IAV-caused mortality as well as morbidity are lowest in children under 5 years of age and highest in mid-aged adults (30-40 years) (6). The age-dependent differences in the clinical outcome of IAV infection are not fully explained by pre-existing comorbidities or maternal transfer of adaptive anti-influenza immunity to infants and point to hitherto unidentified immunological peculiarities that protect infants against IAV. Children have also less severe symptoms when infected with SARS-CoV-2 (7, 8). Several hypotheses for the age-related difference in the severity of coronavirus disease 2019 (COVID-19) are discussed, such as more robust type I interferon (IFN) response, more effective T cell immunity (9), and higher expression of SARS-CoV-2 sensing receptors and inflammatory baseline activation of pediatric compared to adult airway epithelial cells (AECs) (10–12). However, the causes and biological meaning of such differential programming of anti-viral immunity at different ages and in dependency of the type of virus remains elusive.

Apart from the age dependency there is also strong heterogeneity in the course and outcome of RVIs among individuals of same age, pointing to considerable genetic variability or differential development of immunity in concert with their environment. Understanding inter-individual variation and its causes has important implications for targeting patients for escalation of care, inclusion in clinical trials, and individualized medical therapy including vaccination. Besides clear biological differences such as age, sex, race, presence of comorbidities, and genetic variation, differential maturation of immunity during environmental adaptation after birth might result in a different education and regulation of immune responses against RVIs. In humans, the key factors influencing the reprogramming of immunity towards RVIs are still poorly defined and comparison of their effect sizes is long overdue.

A myriad of data has demonstrated that the immune system of the fetus and infant is characterized by a high plasticity, which comes with a high susceptibility to lifelong imprinting effects from environmental cues. Over the past two decades strong evidence accumulated that the composition and function of the human microbiome play a particularly critical role for the development of immunity and overall health (13, 14). Several longitudinal studies revealed aberrant trajectories of upper respiratory microbiota during infancy that are associated with an increased susceptibility to respiratory tract infections. These particularly include delayed and low abundance of *Corynebacterium* and *Dolosigranulum* species and high abundance of *Moraxella*, *Haemophilus* or *Streptococcus* species (15–19). However, our knowledge on how exactly these strains impede respective promote immunity against respiratory viruses remains largely elusive.

Furthermore, the gut microbiota seems to have impact on the development of immunity against RVIs. Though hitherto only shown in mice, gut microbiota-derived short chain fatty acids protected against RSV infections by improving type 1 IFN responses and increasing IFN-stimulated gene expression in lung epithelial cells (20), while the presence of segmented filamentous bacteria protected against IAV, RSV as well as SARS-CoV-2 infection by priming resident alveolar macrophages (21). In humans, only an association between specific gut microbial profiles and RSV disease severity in hospitalized infants has been detected so far (22). However, the molecular pathways of this gut-lung axis and their delicate dependency on age and interactions with environmental exposures (e.g., diet, passive smoke, pollutants) remain unclear. Several clinical trials contributed to an increasing body of evidence that probiotics might be an option to lower the susceptibility to RVIs (23–25); however, many questions on how exactly (e.g., strain dependency), to what extent individualized (e.g., age dependency, host microbial background) and when (e.g., neonatal window) to use such interventions are still open (26–28). Addressing these remaining knowledge gaps could enable modulation of airway and gut microbial communities and their metabolic patterns, presenting a potential therapeutic strategy to prevent or ameliorate severe RVIs.

In summary, at specific developmental stages opposite infectious susceptibility can be observed depending on the causative viral strain. Moreover, different trajectories of immune maturation after birth can lead to strong inter-population differences in the susceptibility to severe RVIs. To understand which specific age- and microbiota-related immunological differences and temporal changes are most relevant across various RVIs, we need a comprehensive view of the longitudinal development of anti-RVI immunity and the impact of and interplay with key factors involved. This requires the establishment of a tailored birth cohort study that investigates longitudinally and in-depth the development of the function of the airway epithelial barrier site and professional innate and adaptive immune cells in newborns against different respiratory viruses. Simultaneously, the study will assess the potential clinical and environmental factors including the status of the respiratory and gut microbiota that may influence these processes. Such comprehensive data sets are mandatory for the development of novel interventions that either target the host or the microbiota to early prevent or successfully treat severe RVIs in the future.

### Aim

The MIAI study hypothesizes that: (i) the course and outcome of RVIs among individuals is related to the large variability in maturational trajectories of anti-viral immunity and (ii) environmental, lifestyle and dietary factors influence the early imprinting of anti-viral immunity in ways that either promote an increased susceptibility to RVIs and their complicated course or support protective patterns (Figure 1). To verify these hypotheses, the MIAI birth cohort has been set up and includes healthy newborn infants born at term and their families. We aim at a comprehensive characterization of age-related differences and changes in the human immune system of the participants with a focus on the function of the cellular key players involved in the establishment of infections with RSV, IAV and SARS-CoV-2. Detailed phenotyping including factors potentially promoting and confounding the individual immune trajectory will be established; in particular, we are interested in the role of the developing microbiome on priming and regulating anti-RVI immunity. The specific aims of our study are:

▪ Aim 1. To characterize the developmental changes of innate and adaptive immune responses to RVIs during the first year of life.
▪ Aim 2. To determine the impact of sociodemographic background, clinical histories and environmental exposures including lifestyle, vaccinations and dietary factors on the trajectories of immune responses toward RVIs.
▪ Aim 3. To elucidate whether there is a link between the microbiome establishment and anti-RVI immunity developing after birth.

**Figure 1.**
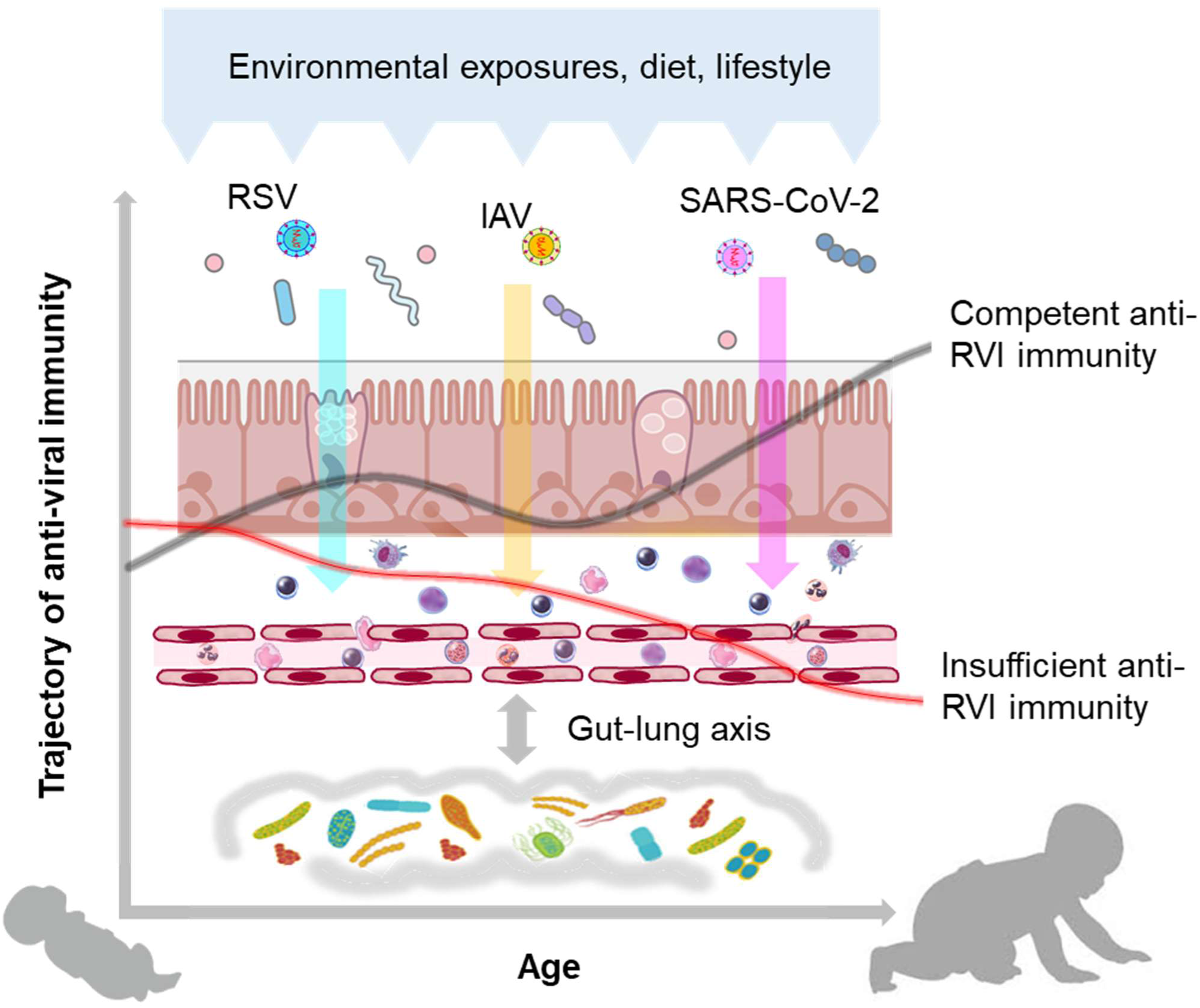
Hypothesis of the MIAI study. The hypothesis of the MIAI study is that the maturation of anti-viral immunity toward RVIs in early life underlies a large developmental variability, which is imprinted by clinical and environmental factors such as dietary habits, lifestyle, and particularly the composition and function of the co-evolving respiratory and gut microbiota.

In brief, we aim to provide insights into age-dependent programming and early-life changes of immunity toward RVIs in humans, highlighting training, priming and acquisition of tolerance, by collecting granular clinical metadata and performing comprehensive unbiased immune phenotyping using multi-omics approaches (Figure 2).

**Figure 2.**
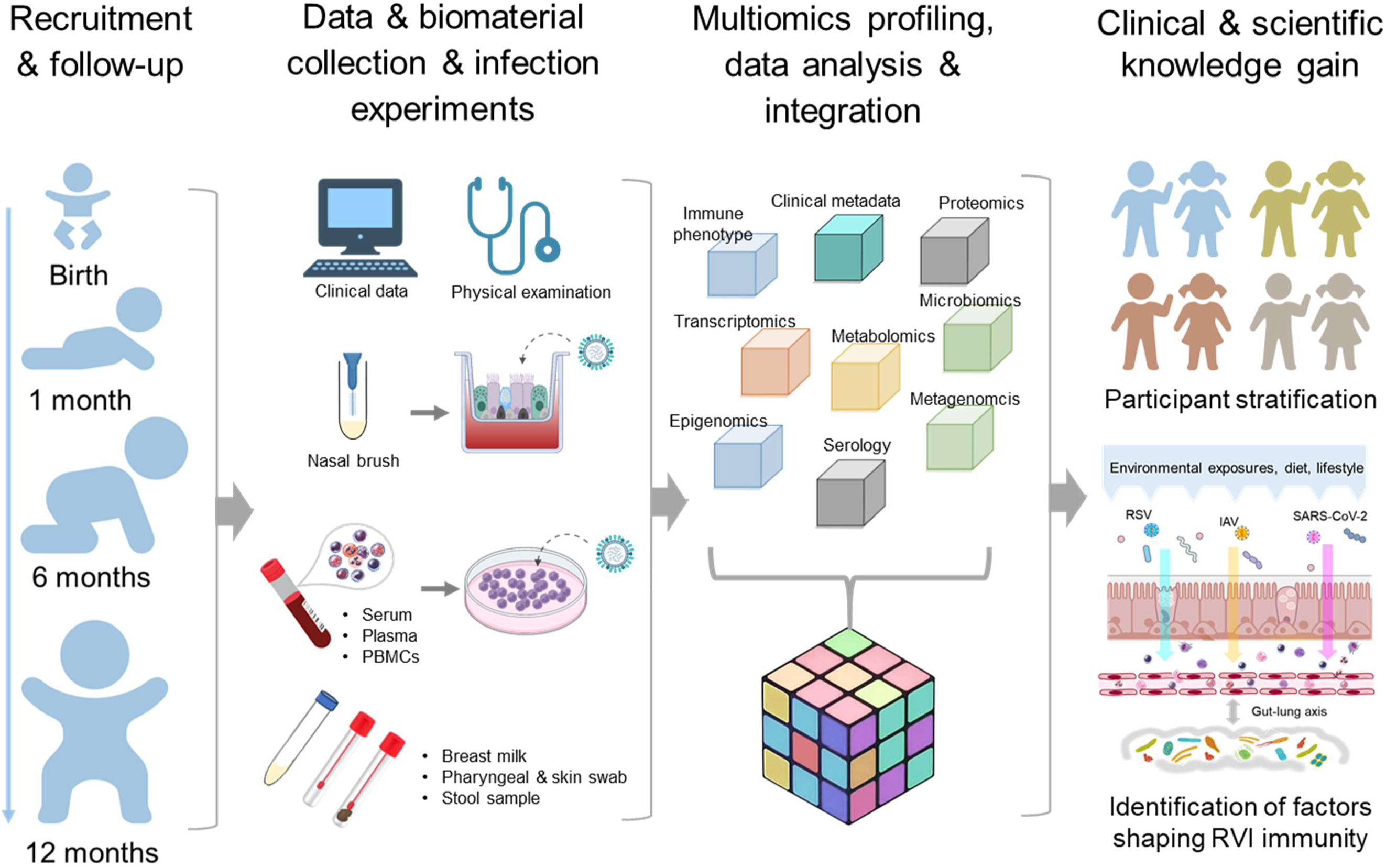
Design MIAI study. The MIAI study encompasses four phases starting with recruitment of term healthy infants and follow-up visits at defined timepoints. At each appointment, clinical data and biomaterials are collected for storage and/or usage in *ex vivo* respiratory viral infection models. Multi-omics datasets will be generated including immune profiles, transcriptomics, epigenomics, proteomics, metabolomics and microbiomics. To gain clinical and scientific knowledge, overall integration of mixed omics data shall enable participant stratification based on marker patterns linking to RVI susceptibility and reveal host and environmental factors imprinting anti-RVI immunity in the first year of life.

## METHODS AND ANALYSIS

### Study design, cohort recruitment and follow-up visits

The MIAI study is a single center, population-based birth cohort study at the University Hospital Würzburg. MIAI enrolls healthy newborn infants and their families and follows them throughout the first year of life (Figure 2). The ongoing recruitment started in February 2022 reaching about 170 infants up to date (with a 2-month interruption of recruitment due to an interim shortage of staff) (Table 1). Inclusion criteria encompass full-term, healthy neonates (37^+0^ to 41^+6^ gestational weeks) born at the perinatal center of the University Hospital Würzburg, Germany, whose familieś residency is located in the city of Würzburg or its administrative district. Exclusion criteria are amniotic infection syndrome, early-onset neonatal sepsis, congenital malformations, primary immunodeficiency or metabolic diseases, and remarkable language barriers. We will however do an earnest attempt to provide understandable information on our study in the native language of parents whose infants would be eligible thus aiming for a population-representative sample. The study recruits a convenience sample cohort; as such, the sample size is not calculated.

**Table 1.** Baseline characteristics of the MIAI cohort.

|  |  |
| --- | --- |
| Total number | 171 |
| Sex, female (%) | 85 (49.7) |
| Gestational age [weeks] (SD) | 39.8 (± 1.2) |
| Mode of delivery<br>VD (%)<br>CS total (%) | 106 (62)<br>65 (38) |
| Multiples<br>Singles (%)<br>Multiples (%) | 160 (96)<br>6 (4) |
| Antenatal corticosteroids | 4 (2.4) |
| Mean umbilical cord arterial pH (SD) | 7.2 (± 0.1) |
| APGAR score<br>Mean APGAR at 1 min (SD)<br>Mean APGAR at 5 min (SD)<br>Mean APGAR at 10 min (SD) | 8.9 (± 0.9)<br>9.7 (± 0.5)<br>9.9 (± 0.3) |
| Premature rupture of membranes (%) | 47 (27) |
| Bonding in the delivery room (%) | 163 (95) |
| Mean birth weight [g] (SD) | 3429 (± 459) |
| Mean percentile birth weight [g] (SD) | 54 (29) |
| Mean birth length [cm] (SD) | 51 (± 2.0) |
| Mean head circumference at birth [cm] (SD) | 34.9 (± 1.4) |
| SGA (%) | 14 (9) |
| LGA (%) | 24 (14) |
| Siblings<br>None (%)<br>One (%)<br>Two or more (%) | 85 (50)<br>67 (39)<br>19 (11) |
| Siblings in child care or school (%) | 74 (43) |
| Mean number household members (SD) | 3.6 (± 0.7) |
| Smoking in family (%) | 37 (22) |
| Pets (%) | 37 (22) |
| Environment |  |
| Rural (%) | 84 (49) |
| Urban (%) | 87 (51) |

SD, standard deviation; CS, cesarean section; VD, vaginal delivery; SGA, small for gestational age; LGA, large for gestational age

The first study visit is performed at the University Hospital for Women’s health and Obstetrics Würzburg. Mothers of eligible infants are approached in the first three days after birth. After obtaining written informed consent from the parents, the baseline assessment takes place during the hospital stay of mother and infant. The follow-up appointments are performed one month, six months and one year after birth at the dedicated Pediatric Clinical Study Outpatient Service at the University Children’s Hospital Würzburg. We are planning to track the study participants for long-term follow-up during childhood.

Overall, the acceptance of our study design with its follow-up appointments and sample collection is high with a relatively low drop-out rate of about 8% after one year as of May 2024, including families that relocated outside of Würzburg and its administrative district. The baseline characteristics of infants enrolled so far are presented in Table 1. Overall, the proportions of female and male infants are equal and the mean gestational age and birth weight appropriate and in line with the inclusion criterium of full-term birth. The largest number of neonates was born vaginally (VD, 62%). Cesarean sections (CS, 38%) were in the majority of CS secondary (60%), i.e. after rupture of membranes, and only a minor part elective (37%) or due to emergency (3%). Half of the MIAI infants have older siblings which of 86% are visiting child care or school. The exposure to nicotine due to smoking family members affects 22% of the MIAI infants and 22% of the families have pets. The distribution of rural compared to urban living environments is even.

Characteristic values of pregnancy histories from infants enrolled in MIAI including maternal factors potentially impacting on the child’s RVI immunity are listed in Table 2. The mean weight gain during pregnancies was 14 kg with a mean body mass index (BMI) of 29.4 (± 5.8) at birth, not exceeding the limits recommended by obstetricians (29). The smaller number of mothers were primigravida (38%). During routine prenatal screenings, gestational diabetes mellitus was detected in 12% and gestational hypertension in 1.8%. The proportions of mothers vaccinated and/or recovered from IAV and SARS-CoV-2 infections are 48% and 97%, respectively. For the Influenza status, 52% of mothers did not know whether they have experienced an IAV infection, while only 3% of mothers did not know their SARS-CoV-2 status.

**Table 2.**
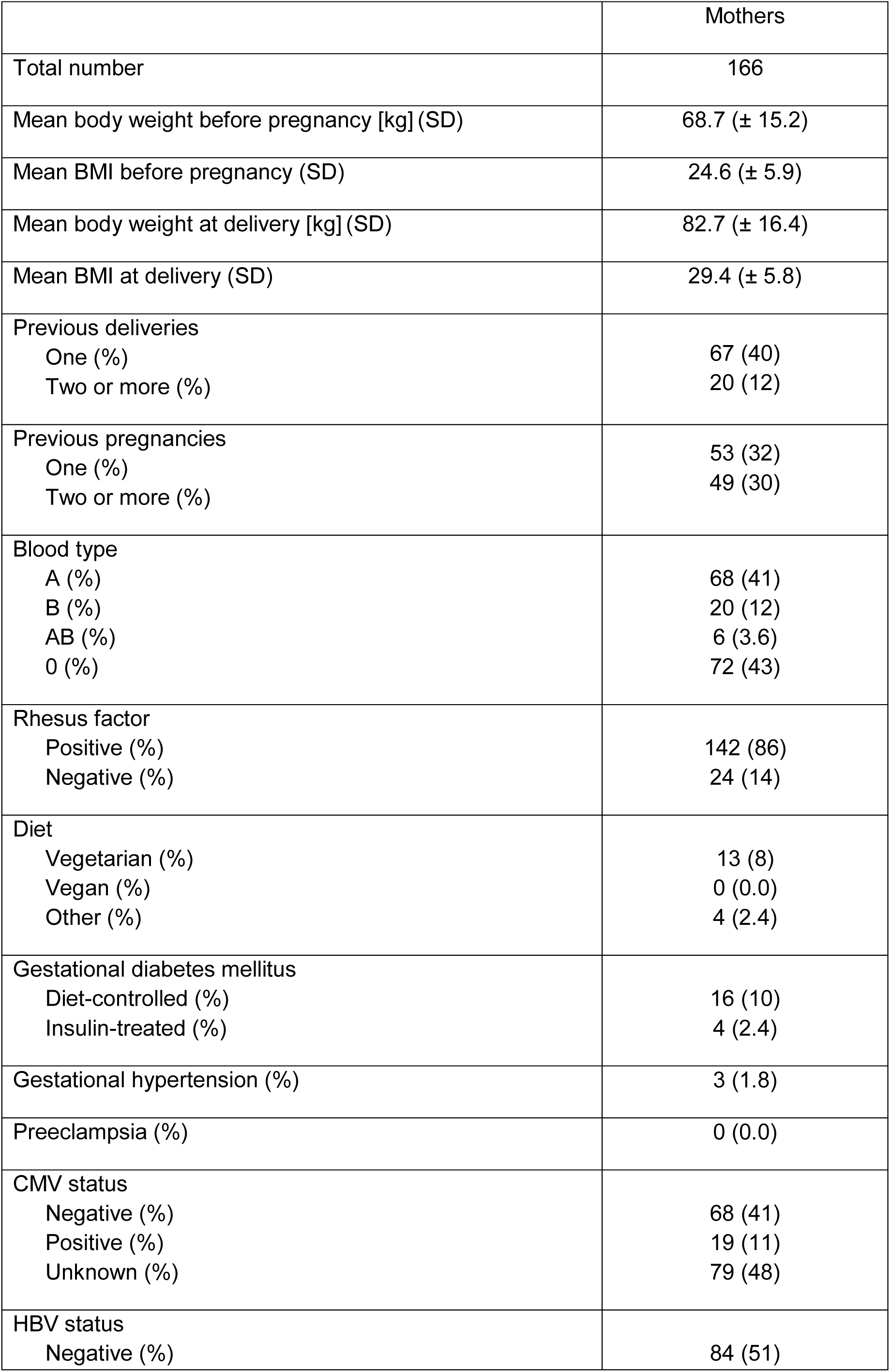
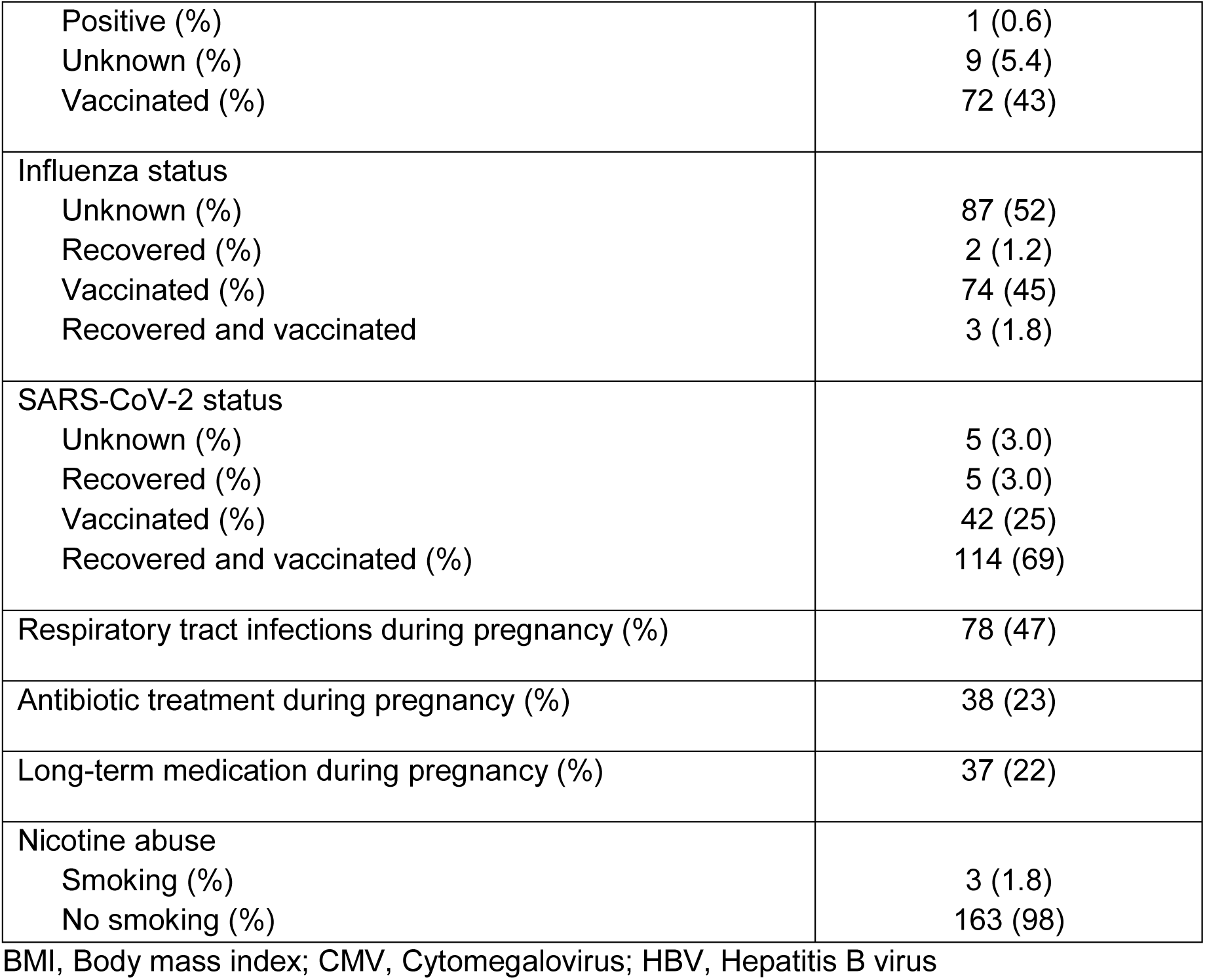
Baseline characteristics of pregnancy in the MIAI cohort.

|  | Mothers |
| --- | --- |
| Total number | 166 |
| Mean body weight before pregnancy [kg] (SD) | 68.7 (± 15.2) |
| Mean BMI before pregnancy (SD) | 24.6 (± 5.9) |
| Mean body weight at delivery [kg] (SD) | 82.7 (± 16.4) |
| Mean BMI at delivery (SD) | 29.4 (± 5.8) |
| Previous deliveries |  |
| One (%) | 67 (40) |
| Two or more (%) | 20 (12) |
| Previous pregnancies |  |
| One (%) | 53 (32) |
| Two or more (%) | 49 (30) |
| Blood type |  |
| A (%) | 68 (41) |
| B (%) | 20 (12) |
| AB (%) | 6 (3.6) |
| O (%) | 72 (43) |
| Rhesus factor |  |
| Positive (%) | 142 (86) |
| Negative (%) | 24 (14) |
| Diet |  |
| Vegetarian (%) | 13 (8) |
| Vegan (%) | 0 (0.0) |
| Other (%) | 4 (2.4) |
| Gestational diabetes mellitus |  |
| Diet-controlled (%) | 16 (10) |
| Insulin-treated (%) | 4 (2.4) |
| Gestational hypertension (%) | 3 (1.8) |
| Preeclampsia (%) | 0 (0.0) |
| CMV status |  |
| Negative (%) | 68 (41) |
| Positive (%) | 19 (11) |
| Unknown (%) | 79 (48) |
| HBV status |  |
| Negative (%) | 84 (51) |
| Positive (%) | 1 (0.6) |
| Unknown (%) | 9 (5.4) |
| Vaccinated (%) | 72 (43) |
| Influenza status |  |
| Unknown (%) | 87 (52) |
| Recovered (%) | 2 (1.2) |
| Vaccinated (%) | 74 (45) |
| Recovered and vaccinated | 3 (1.8) |
| SARS-CoV-2 status |  |
| Unknown (%) | 5 (3.0) |
| Recovered (%) | 5 (3.0) |
| Vaccinated (%) | 42 (25) |
| Recovered and vaccinated (%) | 114 (69) |
| Respiratory tract infections during pregnancy (%) | 78 (47) |
| Antibiotic treatment during pregnancy (%) | 38 (23) |
| Long-term medication during pregnancy (%) | 37 (22) |
| Nicotine abuse |  |
| Smoking (%) | 3 (1.8) |
| No smoking (%) | 163 (98) |

BMI, Body mass index; CMV, Cytomegalovirus; HBV, Hepatitis B virus

Table 3 summarizes anthropometric, medical and sociodemographic characteristics of participating parents. The mean age of mothers and fathers from the entire cohort is 32.8 (± 4.5) years and 35.3 (± 5.4) years, respectively. The medical history data of the parents showed that 52% of the mothers and 24% of the fathers reported somatic disorders including allergies, psoriasis, neurodermatitis, cardiovascular disorders, obesity, thyroid disorders, as well as benign and malignant tumors. Mental disorders were indicated by 8.4% of the mothers and 6.0% of the fathers. About 84% of the participating parents reported a Western European ethnicity and 79% of the mothers and 71% of the fathers were higher educated.

**Table 3.**
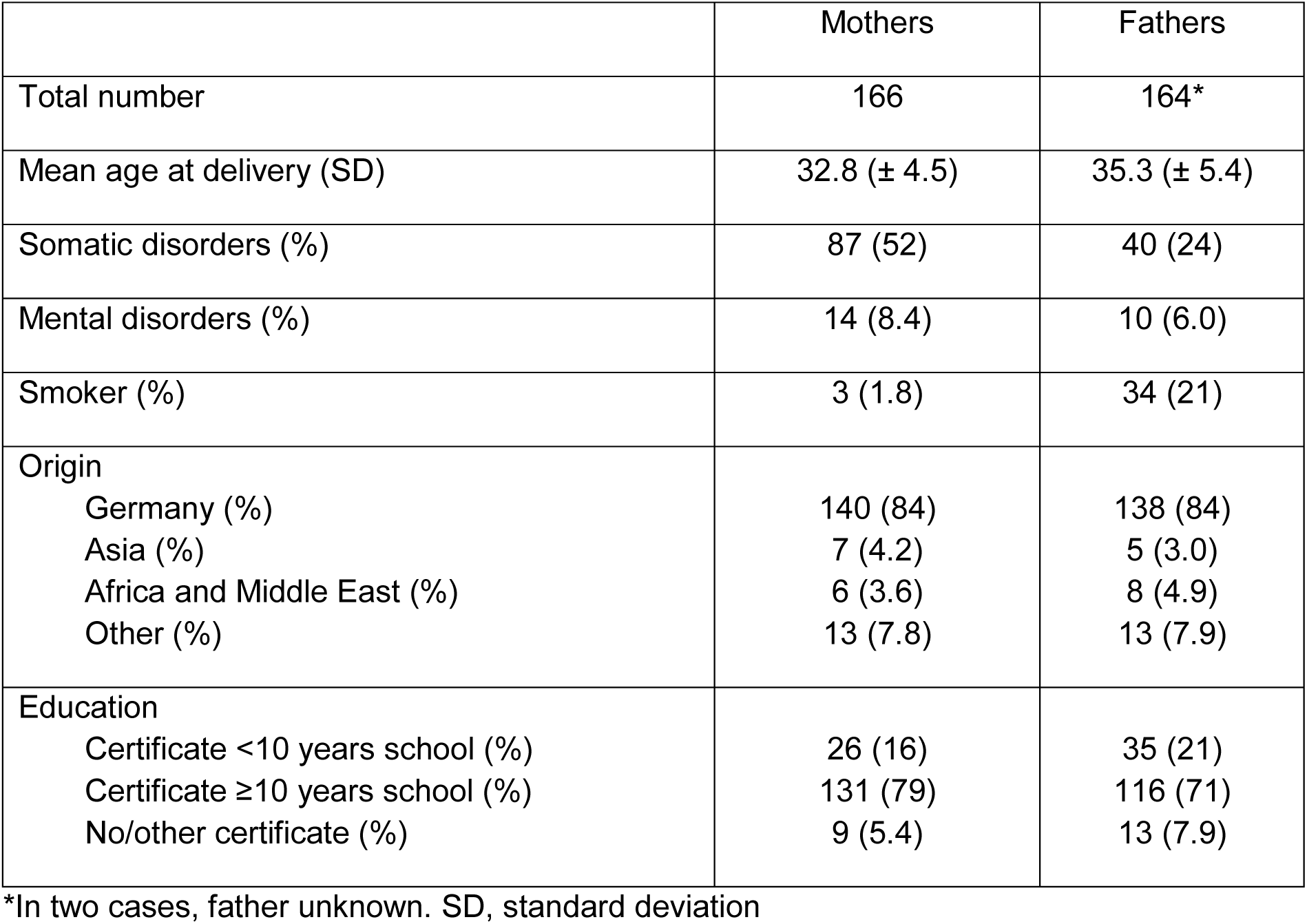
Baseline characteristics of the parents in the MIAI cohort.

### Clinical data collection and data management

Clinical data collection in MIAI is accomplished using tailored case report forms (CRFs) at each study timepoint (Table 4). At baseline, the histories of pregnancy and perinatal events are documented upon enrollment. From mothers and fathers, detailed information on demographic and psychosocial background, family histories and lifestyles are requested. Particular attention is paid on maternal respiratory infections and vaccinations, before and during pregnancy. For the infants, we document at each study visit body measurements, the histories of vaccinations, infections, antibiotics including days of treatment, medications including inhalations, episodes of wheezing, skin rashes, allergies, hospital stays, and surgical procedures, diets including dietary supplements, lifestyle including the number of siblings and further household members, pets, residence (rural/urban), and stays abroad. In addition to the questionnaires, the MIAI pediatrician performs a comprehensive physical examination of the infant at each study appointment to monitor the overall health status including growth, physical and psychomotor development (Table 4).

**Table 4.** Overview of study appointments and clinical data collected in MIAI.

| **Age of the child** | **Day**<br>**1 - 3** | **Day**<br>**30 (-40)** | **Month**<br>**6 (-7)** | **Month**<br>**12 (-14)** |
| --- | --- | --- | --- | --- |
| **Screening for enrollment** | O |  |  |  |
| **Pregnancy and perinatal history**<br>History of diseases during pregnancy including gestational diabetes and hypertension, antenatal and perinatal antibiotics, mode of birth, multiplicity, sex, gestational age, umbilical artery pH and base excess, APGAR, body measurements, complications at delivery | O |  |  |  |
| **Physical examination child**<br>Total body physical examination, anthropometric measurements, blood pressure, respiratory rate, neurodevelopmental milestones | O | O | O | O |
| **CRF child**<br>Body measurements and notes as documented in the child health record book by the attending pediatrician, history of vaccinations, infections, antibiotics including days of treatment, medication, inhalation, episodes of wheezing, skin rashes, allergies, hospital stays, and surgical procedures, diet (exclusive breastfeeding, mixed feeding with formula, age at introduction of solid food) and supplements, day care, lifestyle including further siblings, household members, pets, and residence (rural/urban), stays abroad, family history | O | O | O | O |
| **CRF mother**<br>Age, somatic or psychiatric diseases, previous pregnancies, height, weight before and at end of pregnancy, BMI, CMV/HBV/HIV status, vaccination before and during pregnancy, blood type, infections, medication, diet, smoking, ethnicity, educational level, lifestyle, family history, subjective stress level | O | O | O | O |
| **CRF father**<br>Age, somatic or psychiatric diseases, medication, smoking, ethnicity, educational level, lifestyle, family history | O |  |  |  |

To align with the FAIR data principles (Findable, Accessible, Interoperable, Reusable), the collected clinical data is managed to ensure high standards of data quality and usability. For data protection reasons, the participant’s data is pseudonymized and assigned a unique study ID, which is documented in a separate and password-secured data file accessible by authorized MIAI staff only. A separate external data file is used as a contact database for tracking study participations and invitations to follow-up. The pseudonymized clinical data are double-entered into a password-secured REDCap database hosted at the Institute of Clinical Epidemiology and Biometry, University of Würzburg (IKE-B). Local password-secured copies of the database are generated. The database provides a comprehensive log/audit feature to record data input and changes based on user and time/date. By adhering to these FAIR principles, MIAI enhances the value of its clinical data, promoting its use in further research while maintaining high standards of data security and participant confidentiality.

### Collection and storage of biomaterials

At each appointment, biomaterials from mothers (breast milk) and child (nasal brushes, pharyngeal swabs, skin swabs, stool, blood) are collected according to the schedule illustrated in Figure 3. We have chosen these timepoints to afford serial profiling while minimizing burden. Collection and storage of biomaterials are performed according to established standard operating procedures (SOPs). After collection, biomaterials are immediately transferred to the adjacent laboratory for processing and biobanking.

**Figure 3.**
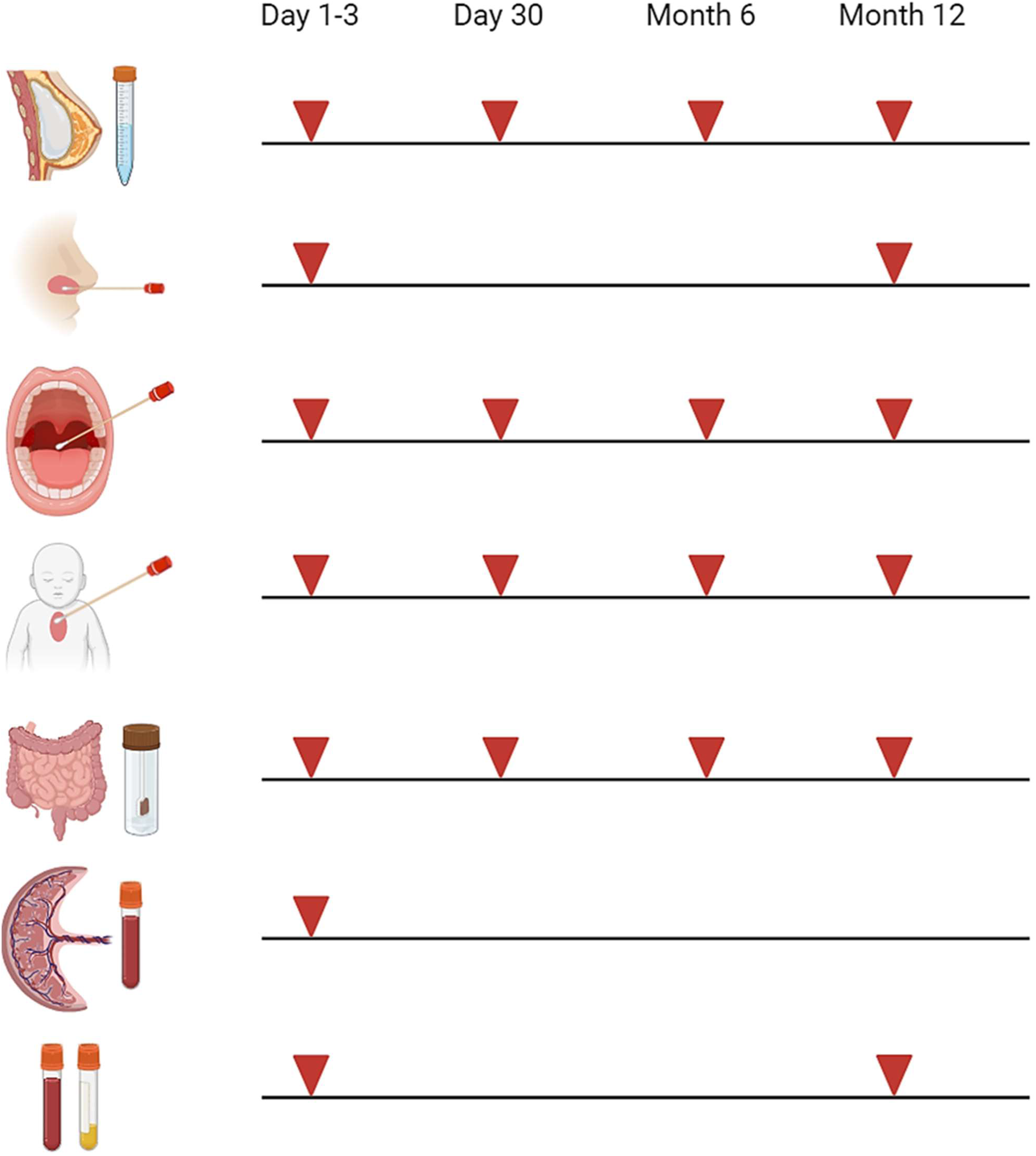
Collection of biomaterials in MIAI. Biomaterials collected in the MIAI cohort include breast milk, nasal brushes and secretions, pharyngeal swabs, skin swabs, stool, cord blood and peripheral blood (EDTA and serum). They are obtained at the defined study time points, i.e. either at day 1-3 and/or day 30 of life and/or at 6 months and/or 1 year following birth. (Created with BioRender.com)

#### Breast milk samples

As long as the mother is breastfeeding, breast milk/colostrum are sampled at each appointment. First, cells are pelleted by centrifugation and the skim milk and the fat layer are removed. The skim milk and fat layer are once more sharply centrifuged and aliquoted for storage at -80°C for later proteomic and microbial studies. The cells are cryopreserved at -152°C following multiple washing steps for later single cell studies.

#### Nasal brushes

Nasal brushing is an established method in pediatric pulmonology, e.g. essential for diagnosing primary ciliary dyskinesia, and provides a robust technique to generate AEC cultures in term and even preterm infants (12, 30). Brushings are taken from the inferior nasal turbinate using sterile interdental brushes (Rossmann, ISO sizes 4 or 5). After sampling, brushes are immediately placed in sterile tubes with transport medium (MEM without additives, antibiotics and antimycotics) and processed within 1 h of collection. Brushes are carefully moved up and down to scrape the cells off and collect them into the transport medium. Next, cells are centrifuged and supernatants stored as “nasal secretions” at -80°C for later proteomic, cytokine and microbiota profiling. The AECs are first expanded in submersion according to established protocols (31, 32) and then cryopreserved at -152°C. Some of the expanded AECs are directly differentiated at the air-liquid-interface (ALI) into a mature pseudostratified respiratory epithelium as described previously (32) and used for infection experiments with RSV (RSV-A-ON1, HRSV/A/DEU/H1/2013) (33) and IAV (H1N1, A/Netherlands/602/2009).

Infections with SARS-CoV-2 (SARS-CoV-2/human/DEU/REGS-200701-CA/2020) are done later under biosafety level 3 conditions using cryopreserved AECs for differentiation at ALI. After infection, AECs are harvested and subjected to immediate FACS studies capturing cell composition and cell quality, generation of total RNA lysates for the analysis of transcriptomes (bulk RNASeq) and viral loads (qRT-PCR), and standardized cryopreservation (34) for later single cell (scRNASeq) and epigenetic (ATACSeq) studies. Apical and basolateral supernatants are stored at -80°C for later analyses of proteome (Mass Spectrometry) and cytokine (FACS-based multiplex assays) profiles and virus plaque assays assessing viral progeny. This approach aims at generating data from each MIAI participant regarding individual airway epithelial immune responsiveness and function toward RVIs at birth and at 1 year of life (Figure 3). The strategy is based on previous reports that ALI-AEC models established from human primary AECs recapitulate well the *in vivo* phenotype of the airway epithelium (12, 35, 36).

#### Pharyngeal and skin swabs

Three pharyngeal swabs and two skin swabs are collected, respectively. One of the respective swabs is transferred to a sterile tube containing sterile PBS with 10% BSA (for protein stabilization) and stored at -80°C for protein and cytokine measurements. The other two respective swabs are stored at - 80°C without additives for later DNA isolation and microbiota studies (16S rRNA sequencing and shotgun metagenomics).

#### Stool samples

Stool samples are collected by the MIAI staff from diapers and immediately transferred to - 80°C. In cases where stool collection at the respective appointments not succeeds, we use samples collected and stored at 4°C by the parents at the evening before or the morning of the follow-up appointment for biobanking at -80°C. Three 1.5 ml tubes are filled with stool, respectively, one of which contains 600 µl DNA/RNA shield (Zymo Research, California, USA) to preserve nucleic acids for 16S rRNA sequencing and metagenomics studies. The other two stool aliquots are stored without additives and used for the extraction and analysis of immunoactive proteins such as S100-alarmins (37) and cytokine profiles according to established protocols (38).

#### Blood samples

Cord blood is collected preemptively from all eligible neonates but only processed when parents agree on participation in the MIAI study. If no consent is given, cord blood samples are discarded. The volumes of drawn peripheral venous blood samples align to the pediatric guidelines of the European Medical Agency, which recommends that blood collection for research purposes should be at maximum 1% of total blood volume (i.e., 0.8 ml/kg body weight). At the first sampling timepoint, we seek to combine blood collection in MIAI with the routine newborn screening taken between 36 and 72 hours of life. Blood samples are processed within 1 h after collection. The serum samples are stored at -80 °C and used for cytokine, metabolome and serological studies including quantification of antibody titers against RSV, IAV and SARS-CoV-2. From EDTA samples, peripheral blood mononuclear cells (PBMCs) are isolated and profiled using established multidimensional FACS panels (39–43). One part of isolated PBMCs is cryopreserved at -152°C for later single cell, epigenetic and experimental studies (e.g., analysis of T and B cell subsets and receptor repertoires including virus-specific effector T cells). To avoid functional impairment of the myeloid cells due to cryopreservation, monocytes are directly isolated from the other part of isolated PBMCs as described previously (44, 45) and infected with RSV and IAV. After infection, total RNA lysates are generated and culture supernatants harvested and both stored at -80°C for the analysis of transcriptomes, viral loads, proteome and cytokine profiles, and viral progeny.

### Data analysis and statistics

Omics data analysis in MIAI will be performed on different levels as results from transcriptomics including lymphocyte receptor repertoires, epigenomics, 16S rRNA gene and shot-gun metagenomics sequencing, breast milk and serum metabolomics and proteomics including cytokine and serologic profiles. We aim at subjecting full longitudinal sample series from 400 infants to study the omics profiles. Given the longitudinal nature of our study set up, samples previewed for these studies will be randomized to reduce batch effects. Initially, each omics dataset will undergo individual analysis to elucidate its unique characteristics. Subsequent exploratory analyses will determine the necessity of batch correction for each dataset. If required, corrections will be implemented using ComBat batch method to normalize across multiple analytical runs and WithinVariation function implemented in the R mixOmics package and to normalize features as change over time within each child. Post-correction, we will validate that the biological variability is preserved and that no artifacts were introduced. We will perform omics-specific statistical analyses and co-expression network analyses using hCoCena (46, 47) to identify and characterize differential subgroups with greater precision. All data generated including standardized meta-data will be quality-controlled and gathered within a unified framework following standards for omics and clinical measurement representation, allowing within-study integration and comparative analysis with previous studies according to FAIR data principles.

Differential trajectorial endotypes will be identified using KmL (k-means for longitudinal data) clustering. To determine the optimal number of clusters (k), we will run 1000 permutations, utilizing the Harabasz criterion (Calinski-Harabasz index) to evaluate clustering validity. This methodology will facilitate a nuanced, continuous classification of patients, transcending binary categorization and allowing also for a more detailed representation of endotypic variation. Specialized software packages for dataset-specific imputation will be employed to address any missing data points, ensuring robust classification of immune and microbiome trajectories across the cohort. Advanced statistical methods and machine learning algorithms will be integrated into the analysis pipeline to enhance the precision and reliability of the endotype classification.

To elucidate the relationships between specific immunotypes, respiratory or gut microbiome profiles, environmental factors, and clinical outcomes, we have created two specialized software tools, metadeconfoundR (48) and longdat (49). These tools enable comprehensive mixed omics data analysis while managing the complex interplay of numerous demographic, therapeutic, and disease-related variables. MetadeconfoundR is designed to facilitate the identification of biomarkers within cross-sectional multi-omics medical datasets, while carefully accounting for confounders. Initially, it identifies significant associations between features and metadata using nonparametric tests such as the Mann-Whitney U test. Subsequently, it assesses and adjusts for confounding effects among these metadata variables through *post-hoc* nested linear model comparison. Importantly, metadeconfoundR can integrate previously established knowledge of confounders, such as drug associations from the MetaCardis cohort, into its analysis. This inclusion aids in maintaining robust conclusions even when new datasets lack the statistical power to independently identify these confounders. LongDat, meanwhile, is an R package tailored for analyzing longitudinal multivariable data, effectively handling a wide array of covariates. It distinguishes direct from indirect intervention effects and identifies key covariates that may act as mechanistic intermediates. Although primarily focused on longitudinal microbiome data, LongDat’s versatility allows application to various data types including binary, categorical, and continuous data. Comparative testing against other analytical tools like MaAsLin2, ANCOM, lgpr, and ZIBR has demonstrated LongDat’s superior performance in terms of accuracy, runtime, and memory efficiency, making it an ideal choice for high-dimensional longitudinal studies where multiple covariates are a consideration.

### Next steps and outlook

To gain a comprehensive picture of how immune profile development including cellular responses to RVIs varies amongst individuals, we are in the process to analyze the biomaterials by a multi-omics approach. This encompasses transcriptomic, epigenomic, metagenomic, and metabolomic layers. These data are extended by cellular phenotyping using high-dimensional flow cytometry and the innate and adaptive immune responses to IAV, RSV and SARS-CoV-2. Next, the omic phenotypes will be integrated with large clinical data sets. As most of the one-year-old infants have not yet attended a day care center, which is an important contributor to microbiome maturation (50, 51), we continue the longitudinal assessment of MIAI participants by the means of annual questionnaires until regular follow-up at early school age. Our long-term vision is to disentangle the role of early programming of anti-viral immunity for the susceptibility to RVIs and also non-communicable diseases such as asthma, obesity or autoimmune disorders. Future plans include data collection of participants until 12 years of participants’ age, which would allow to investigate how differences in omics immune and microbiome endotypes translate into population differences in clinical outcomes and susceptibility to RVIs.

## DISCUSSION

Considering the impact of the recent SARS-CoV-2 pandemic (52) or previous pandemics and epidemics due to IAV and RSV on human health (6, 53, 54), our knowledge about the ontogeny of immunity towards RVIs and the factors determining the developmental variability is little. Except for pre-existing co-morbidities and primary immunodeficiencies, hitherto, only the acquisition of autoantibodies against type I IFNs (55) and genetic variants, particularly in cytokines and pattern recognition receptors (56), could be linked to severe RVI cases, though only in a minor proportion of RVIs, and explain inter-population variability of anti-RVI immunity. We recently initiated the recruitment of a birth cohort of healthy newborn infants in Germany that has been designed to investigate in particular the development of immunity towards RVIs in early childhood. The data shown in the present manuscript reflect an initial characterization of the MIAI cohort. Comparison of the structure of the current MIAI cohort with those reported from other recruiting maternal or term infant birth cohorts in Europe show similar distributions and rates of e.g. sex, gestational age, birth weight, multiple births, mode of delivery, first-time mothers, number of siblings, maternal age and BMI, gestational diabetes and hypertension, infections during pregnancy, and parental education and smoking (57–60). It is gratifying that in MIAI the drop-out rate of 8% is low in comparison to other birth cohorts which also perform repeated collection of biomaterials, e.g., the SweMaMi cohort (59) and KUNO-Kids cohort (57) that recorded 40-50% of lost 1-year follow-ups. Hence, we assume that the MIAI cohort will be representative of an infant population in Central Europe.

The common immunological finding in severe IAV, RSV and SARS-CoV-2 related infectious diseases is an exceeding cytokine release (61–64), suggesting a strong involvement of a systemic inflammatory response due to a lack of viral clearance at the epithelial barrier site or its breakage. Therefore, our first research focus in MIAI will lay on the age-dependent immunological profiling of the airway epithelium and blood monocytes, as key mediators of systemic cytokine storms during severe RVIs (65, 66), and the identification of demographic, clinical, environmental, lifestyle and dietary factors on the trajectories of their immune responsiveness toward RVIs. Interestingly, a previous study revealed a link between a low susceptibility of monocytes to IAV infections and high basal monocyte activation, which the authors assumed to be driven by environmental factors and/or weak-effect genetic variants that remain to be identified (67).

A plethora of studies report on distinct gut and respiratory microbiota compositions that are associated with an increased risk of RVIs with either IAV, RSV or SARS-CoV-2 (13, 15, 16, 18–20, 22, 68–70). How exactly such reported microbiota profiles influence anti-RVI immunity remains largely elusive. To what extent the developmental changes of the microbiota shape the postnatal ontogeny of the development of anti-RVI immunity is even less understood. The MIAI study will help to characterize differential trajectories of anti-RVI immunity and improve our knowledge about influencing factors, in particular the influence of the gut and respiratory microbiota.

The strength of the MIAI study is the population-based approach and the standardized in-depth longitudinal characterization of healthy individuals at birth and the comprehensive analysis of preserved biomaterials. The usage of cutting-edge multi-omics technology for the generation of longitudinal immune-microbiome profiles combined with functional profiling of the cellular immune response against RVIs make this cohort study a unique approach. Finally, by ensuring that the multi-omics-based profiles are integrated with highly granular clinical information, we are confident to identify factors that either promote or impede the development of immunity against RVIs. As in other population-based studies, we observe a bias toward high socioeconomic status among MIAI participants. Technically, the study has its limitation in the standardization of stool sample collection as sometimes home sampling is required, which precludes immediate freezing at -80°C as warranted by on-site visit sampling through the staff. With respect to clinical information, the study would benefit from an app-based monitoring that allows immediate reporting of acute infections that in case of respiratory symptoms then would prompt viral diagnostics. We aim at a timely implementation of this important add-on.

In conclusion, the MIAI birth cohort study expects to generate data that give a holistic picture on the variability of the maturation of immunity towards RVIs in early life and identify host and environmental factors that influence the trajectory. By linking differential trajectories along with clinical information on incidences and outcomes of RVIs to specific immune features, MIAI will pave the way for in-depth elucidation of molecular mechanisms, which is the basis for the development of age-specific host-directed treatment strategies against severe courses of RVIs (e.g., (anti-)cytokine biologicals, tailored cell- or RNA-technology-based therapies). Of particular importance will be the identification of factors shaping anti-RVI immunity, as this is essential to design personalized interventions such as metabolic, probiotic and/or dietary measures that promote developing competent anti-RVI immunity in early life thus preventing severe RVIs in the long term.

## Data Availability

All data produced in the present study are available upon reasonable request to the authors

## ETHICS AND DISSEMINATION

This study was reviewed and approved by the Institutional Review Boards of the University of Würzburg (no. 13/22_z-am). The study is also registered on the German Clinical Trials Register (DRKS). The MIAI cohort is being conducted in compliance with all pertinent legislation and directives and following the guidelines on human biobanks for research and other relevant directives on research ethics. All parents have before inclusion signed informed consent in accordance with the Declaration of Helsinki II. Special emphasis is placed on privacy and safety of data.

The proposed research is anticipated to generate a significant body of knowledge of interest to a wide range of stakeholders, from the clinical (pediatricians and other health professionals) to the research community but also to the public. Hence, a multilayered dissemination strategy will be followed and builds on cross-media communication to reach the specific target groups through their preferred media and information platforms. Local, national and international meetings and publication in peer-reviewed medical journals and via websites are used to inform for the scientific community. Once a year, study participants and healthcare professionals are invited by the study team to join online video meetings where they can discuss medical questions and pose any questions or comments on the study procedures. In addition, participating families will be provided with a copy of this publication to disseminate the MIAI study results.

## Acknowledgements

We would like to extend our gratitude towards families including the mothers, fathers and their infants included in the study, the sample collection team and recruitment team. This study would not have been possible without the work of a large group of staff members from the different departments. In particular, we acknowledge the contributions of the student assistants Lucia Baumgartner, Theresa Wurm and Leon Fagner, the entire team of midwives and the maternity unit including the lactation specialists.

## Author contributions

Study design: MW, SP, CH, DV. Project management: CRH, CH, DV. Recruitment: RK, CRH, MK, EF, JF, MW, JB, CV, MM, JM, AW, DV. Biobanking: JS, MR. Data management: RK, CRH, MW, SP, MK, EF, JF, CV, MM, JM, TK, PUH, DV. Experiments: JS, MR, MW, JH, JD, HM, SS, MS, SH, TP, SB, LD, Sagar, TU, SKF-S. Data analysis: CRH, MW, SP, JH, JD, HM, SB, UL, Sagar, TU, SKF-S, DV. Data interpretation: CRH, MW, SP, JH, JD, HM, MS, SH, TP, SB, LD, UL, Sagar, TU, SKF-S, CH, DV. Writing: RK, UL, TU, CH, DV. Editing and revising: all authors. Funding acquisition: CH, DV.

## Competing interests

The other authors declare no competing interests.

## Funding

This work is supported by grants from Deutsche Forschungsgemeinschaft (DFG, German Research Foundation) to DV (VI 538-9-1), CH (HA-6409-5-1) and TU (UL 521/1-1). The following other funding sources support the project: the Federal Ministry of Education and Research (BMBF) to SP, CH and DV (PROSPER; 01EK2103B and 01EK2103A, respectively); the DFG to SP (PI 1512/1-3) and DV (VI 538/6-3); the DFG SFB 1583/1 (“DECIDE”) project number 492620490 to DV and SB; the DFG TRR 359 (“PILOT”) project number 491676693 to DV; the DFG Germany’s Excellence Strategy – EXC 2155 ‘RESIST’ – Project ID 390874280 to DV and TP; the BMBF Advanced Clinician Scientist Program (“INTERACT”, project number 01EO2108) to HM.

## Notes

### Competing Interest Statement

The authors have declared no competing interest.

### Author Declarations

Ethics committee of the University of Wuerzburg gave ethical approval for this work.

